# Trends in diversity of scientific authorship by ethnicity, sex and age of authors in a National Institute for Health and Care Research Biomedical Research Centre in England: A retrospective longitudinal analysis

**DOI:** 10.64898/2025.12.19.25342411

**Authors:** Lorna R Henderson, F. Eugenio Barrio, David Saunders, Owen Coxall, Vasiliki Kiparoglou, Evanthia Kalpazidou Schmidt, Syed Ghulam Sarwar Shah

**Author notes:** Corresponding author: Syed Ghulam Sarwar Shah.

## Abstract

**Objective:** Scientific publications are important markers of achievement in academic medicine. Whilst gender disparities in scientific authorship are well documented, there is little understanding of the influence of authors’ protected characteristics on authorship positions, publication outputs and citations in biomedical and translational research. This study examined the diversity of scientific authorship by three protected characteristics, i.e., ethnicity, sex and age of authors of biomedical research publications.

**Design:** A retrospective longitudinal descriptive analysis of 476 unique authors’ protected characteristics, biomedical scientific publications and citations from 2012 to 2022.

**Setting:** National Institute for Health and Care Research (NIHR) Oxford Biomedical Research Centre (BRC) in England.

**Data:** Peer-reviewed publications funded by the NIHR Oxford BRC during its funding round 2 (2012–2017) (henceforth BRC2) and round 3 (2017-2022) (henceforth BRC3). BRC author-level data were extracted on three self-reported protected characteristics, i.e., ethnicity, sex (male and female) and age of authors. Data were analysed by descriptive statistics, Fisher’s exact test and Pearson’s chi-squared test of independence to explore the proportions of first and last (senior) authorship positions and number of publications and citations by ethnicity, legal sex and age of authors.

**Primary outcome measures:** Authorship positions (first and last authors), number of publications and citations of publications by author ethnicity, sex and age.

**Results:** There were statistically significant differences in first and last authorship positions and number of publications and citations of publications by author ethnicity, sex and age. White researchers were 79% and 85% of first authors and 85% and 86% of last authors in BRC2 and BRC3, respectively. In BRC2, females were more (53%) in the first author category, while males were more (58%) in the last author category (p<0.014). In BRC3, male first (70%) and last authors (78%) were statistically significantly more than female first (30%) and last (22%) authors (p<0.001). For the total study period, 44% of first and 24% of last authors were female, which was statistically significantly less than male first (56%) and last (76%) authors (p<0.001). Authors aged 40-59 were statistically significantly more (79% and 71% of first authors and 73% and 75% of last authors in BRC2 and BRC3, respectively) than authors of other age groups (p<0.001).

White authors published more publications (87%) than BAME authors (13%) in both BRC periods, and there were no differences in the overall proportions of publications by authors of either ethnicity in both BRC periods (p=0.91). In BRC2 and BRC3, male authors produced statistically significantly more publications (59% and 73%) than female authors (41% and 27%, respectively, p<0.001). Authors aged 40-59 years published more papers (64% and 69% of total papers in BRC2 and BRC3, respectively) than authors of other age groups (p<0.001).

Citations of publications by white authors were statistically significantly higher (89% and 87% of total citations of BRC2 and BRC3 publications, respectively) than publications by BAME authors (p<0.001). Publications by male authors had higher citations (65% and 68% of total citations of BRC2 and BRC3 publications, respectively) than female authors’ publications (p<0.001). Citations of publications by authors aged 40-59 years were higher (56% and 72% of total citations of BRC2 and BRC3 publications, respectively) than citations of publications by authors aged less than 40 years and those aged 60 and above (p<0.001). Authors aged 60-64 years had the highest average citations per publication in both BRCs and the total period of the study. In the BRC3 period, some positive trends were observed, such as a substantial increase in the absolute number of BAME and female first and last authors and young first authors (aged up to 39 years); an increase in the number of publications by BAME, female and young authors (aged up to 39 years); and an increase in total citations of publications by BAME, female and young authors, and higher average citations of female authors’ publications (42 citations per publication) compared to male authors’ publications (33 citations per publication) in BRC3.

**Conclusion:** There are statistically significant differences in diversity of authorship positions, publication output and citations by ethnicity, sex and age of authors. Positive trends include an increase in BAME, female and young authors and their publications and citations; however, under-representation of authors of BAME background, female and younger age is identified. Gaps in scientific authorship, publications and citations by author ethnicity, sex, and age could be addressed by systematic collection of data on protected characteristics of authors (researchers). In addition, tackling disparities in recruitment, career progression and research funding is imperative.

**STRENGTHS AND LIMITATIONS OF THIS STUDY:**

- This is the first study to examine diversity at author level by ethnicity, sex and age to our knowledge in three publication metrics: authorship, publications output and citations count within the context of an NIHR BRC.
- Most studies have examined gender individually, here we explore the influence of ethnicity, sex and age of authors on authorship type (first and last authors), publications productivity and citations count.
- This study provides benchmarking data on scientific authorship diversity of relevance to other NIHR funded BRCs.
- This study identified the challenges of benchmarking protected characteristics at individual author level because such data is not routinely available or collected.
- Completion of protected characteristics data is self-reported, not mandatory and therefore incomplete.
- However, the data does provide important benchmarking data and the feasibility of gathering such information at author level. Further work could be completed to improve the reliability and comprehensiveness of such data.

## INTRODUCTION

Biomedical Research Centres (BRCs) are translational research organisations (TRO’s) established in England in 2007 by the National Institute for Health and Care Research (NIHR) (1). BRCs are partnerships between leading National Health Service (NHS) hospital trusts and universities and they are required to track equality, diversity and inclusion (EDI) improvements as a prerequisite for translational research funding (2,3). Peer-reviewed scientific publications are markers of achievement and determinants of career progression (4,5) Citations are also important markers of success, impact and quality (6–8). Contractually, the number of peer-reviewed publications by BRC-supported authors is reported to the funder i.e. the NIHR (9); however, little is understood about the diversity of the authorship.

Authorship position is an important marker of achievement, with “first authorship” and “last authorship” being the most highly regarded. First authorship denotes the author has completed the majority of the work and it is an important marker to enhance career development e.g. grant applications, fellowships etc (10). Last (senior) authorship position is typically given to senior academics, leads, and group leads and it is an important marker of achievement and seen to demonstrate as good group leadership and funding oversight (11).

### Study objectives

In this longitudinal analysis of peer-reviewed biomedical research publications published over ten years covering two BRC funding rounds: BRC2 (2012–2017) and BRC3 (2017-2022), we examined the influence of three protected characteristics i.e., ethnicity, sex, and age of authors on authorship positions (first and last authors), publication output and citations count. Previous research has examined these factors individually such as gender equity in authorship (9,12), or ethnicity and authorship (13,14) but in this study we examined three variables: ethnicity, sex, and age, together in relation to authorships categories, publication output (productivity) and citations count.

The aim of the study was to benchmark our researchers’ outputs by three protected characteristics i.e., ethnicity, sex, and age and provide an important evidence base on the EDI of researchers in biomedical research in our BRC funded by the NIHR.

## METHODS

### Study design

We conducted a retrospective longitudinal descriptive analysis of peer-reviewed biomedical research publications published between 2012 and 2022.

### Setting

This study was conducted at the NIHR Oxford BRC, which is a research collaboration between the Oxford University Hospitals NHS Foundation Trust and the University of Oxford (15). The NIHR BRCs support translational research and innovation to improve healthcare for patients (16). Our BRC was one of the first BRCs funded by the NIHR in 2007. In 2006-07, the NIHR Oxford BRC was awarded £96m to support research across nine research themes, five cross-cutting themes and a range of underpinning platforms. In 2016, our BRC was awarded £113m funding for five years (2017-2022) to support research in twenty research themes (15).

### Data

Data comprised peer-reviewed publications published by 476 researchers affiliated with the NIHR Oxford BRC. Data were compared in two different BRC funding rounds i.e. BRC2 (2012–2017) and BRC3 (2017-2022). BRC author level data included three self-reported protected characteristics: author ethnicity (white and BAME), legal sex (male/female), and age (in five-year groups). BRC funded research authors are recorded on Symplectic Elements (‘Elements’), the University of Oxford’s current research information system (CRIS). The Elements database holds records of researchers’ publications and other research activity and can report by researchers’ BRC research theme for the purposes of annual reporting to the funder NIHR, which is a contractual requirement.

The eligibility criteria included researcher recorded in Elements as member of BRC theme(s) and at least one peer-reviewed journal article linked to their account in Elements.

Data extracts of 476 distinct BRC researchers, and their affiliation to themes, publication lists (including full bibliographic and bibliometric metadata), and authorship relationship (first and last authors) were downloaded from the Elements database. The team at Research Services responsible for the Elements database liaised with the University of Oxford Human Resources (HR) Department who independently extracted researchers’ three self-reported protected characteristics: i.e., legal sex (male/female), ethnicity (white and BAME - Black and Asian Minority Ethnic) and age from the HR database (CoreHR) of the University of Oxford. Data were provided in aggregate form and anonymously to ensure individual researchers were not identifiable. The process of data capture, linking and analysis is illustrated in Figure 1.

**Figure 1.**
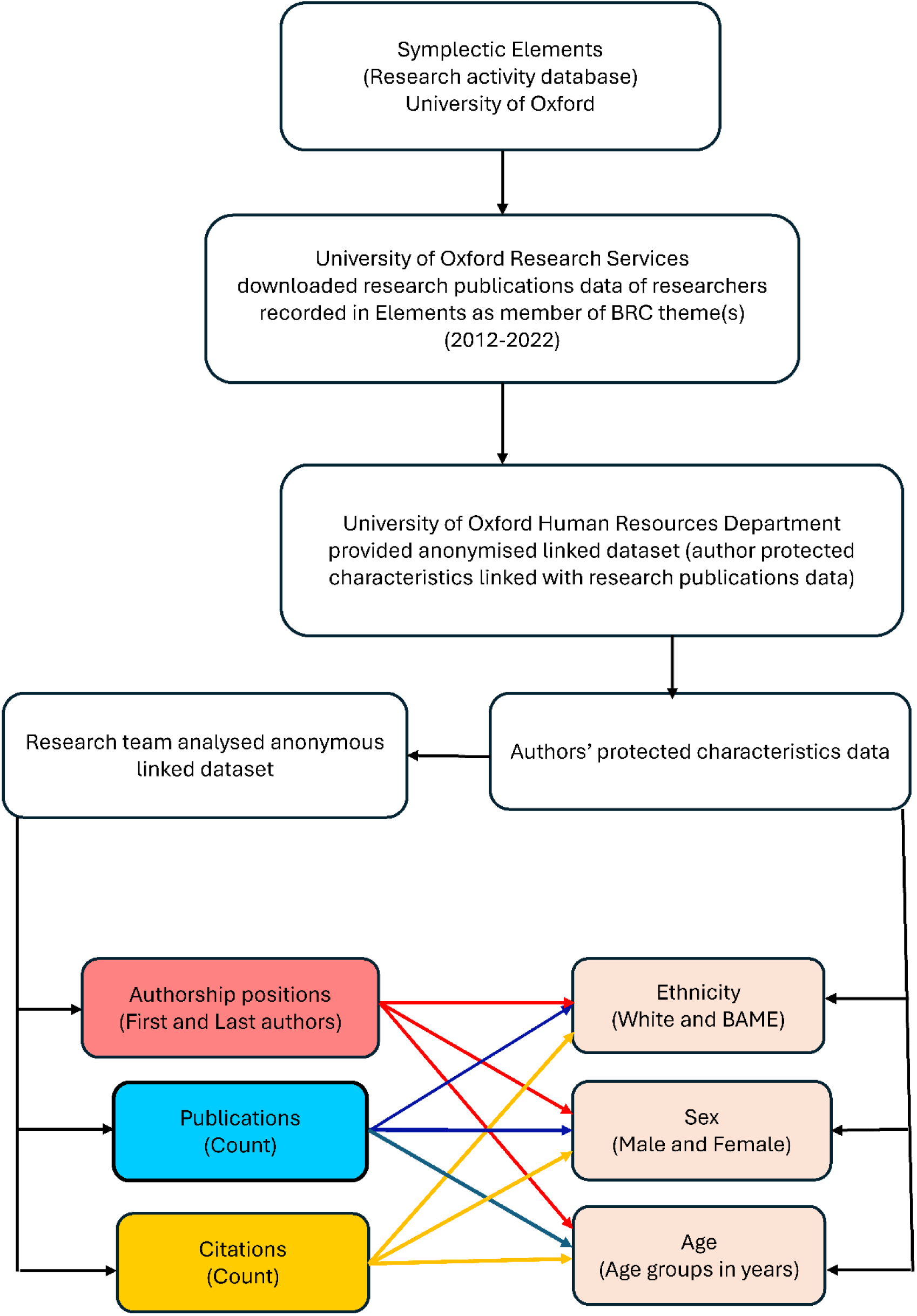
Data capture, linking and analysis process

### Outcome measures

The key outcome measures were three publication metrics: authorship position (first and last authors), number of publications and citations of publications by three protected characteristics of authors: ethnicity (white and BAME), sex (male and female) and age (five-year age groups).

### Statistical analysis

Data were analysed by descriptive statistics, Fisher’s exact test and Pearson’s chi-square test of independence using SPSS, version 28 for windows. In addition, percentage changes in publications and citations in BRC3 compared to BRC2 by author ethnicity, sex, and age were calculated using Microsoft Excel®, which was also used for creating visualisations.

We calculated percentage changes in publications and citations in BRC3 compared to BRC2 by using the following formula:

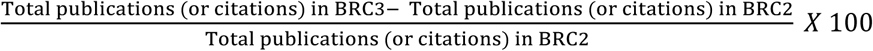

We also calculated average citations per publication by author ethnicity, sex and age by a formula as under:

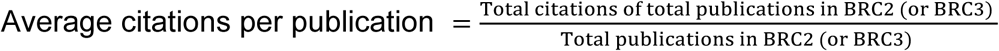

### Patient and public involvement

There was no patient or public involvement in the study design.

## RESULTS

### Authorship position by ethnicity, sex and age of authors

Proportions of authorship positions by ethnicity, sex and age of authors are presented in Table 1. Proportions of white researchers were higher in both first and last authorship positions compared to BAME authors in both BRC2 and BRC3. There were no statistically significant differences in the total proportions of white and BAME first and last authors between BRC2 and BRC3 as well as for the total study period (combined BRC2 and BRC3 periods = 2012-2022) (Table 1).

**Table 1.**
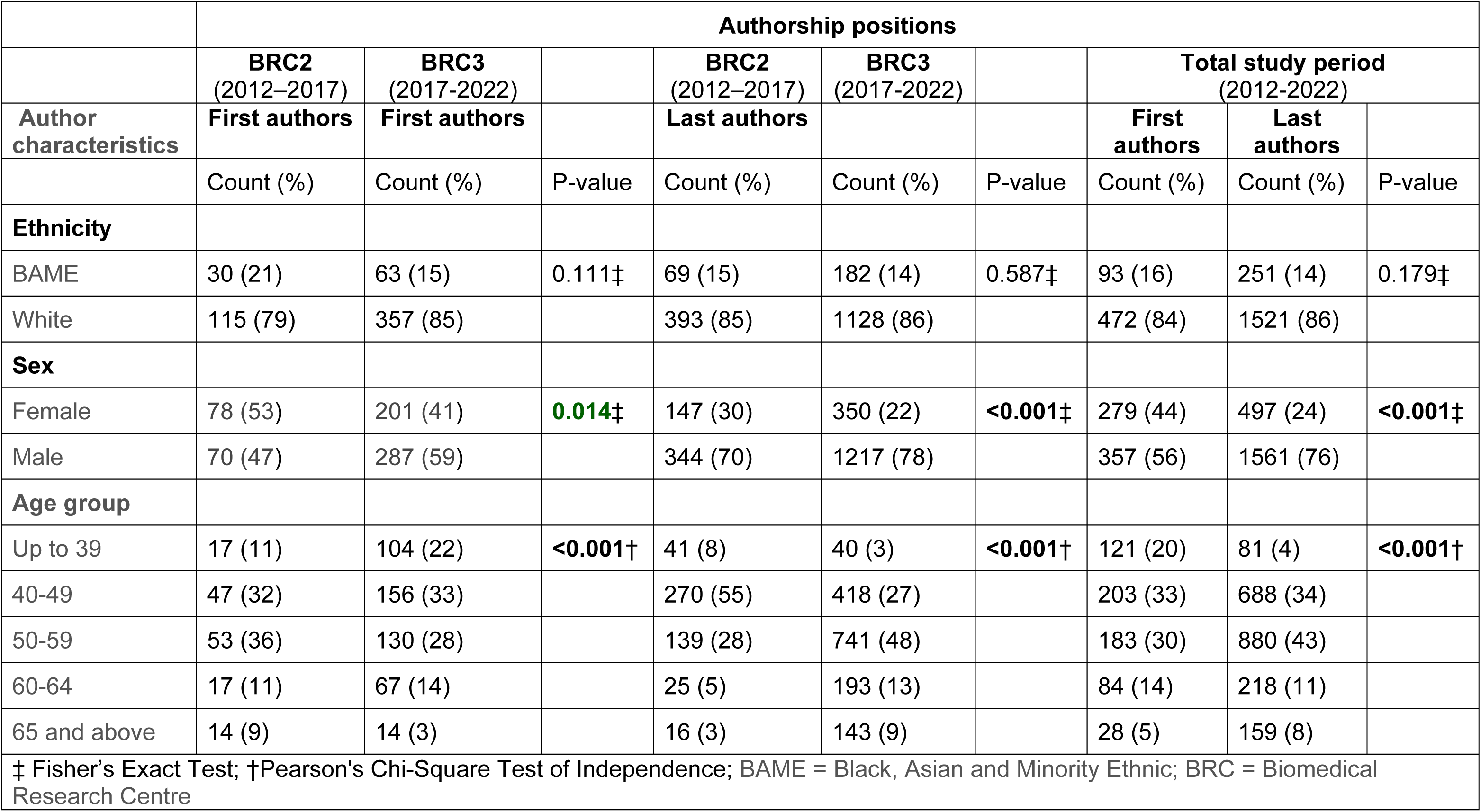
Authorship positions by ethnicity, sex and age of authors.

There were statistically significant disparities in authorship positions by author sex (male and female) (Table 1). Female first authors’ proportion of was higher than male first authors’ proportion in BRC2 but female first authors’ proportion decreased and was lower than male first authors’ proportion in BRC3 (p=0.014). In the last authorship category, male authors’ proportion was statistically significantly greater than female authors’ proportion in both BRC2 and BRC3 (p<0.001). In the total study period, proportions of female first (44%) and last (24%) authors were statistically significantly lower than male first (56%) and last (76%) authors (p<0.001).

Authorship positions by author age (Table 1) also showed statistically significant disparities in proportions of first and last authors of different age groups (p<0.001). In BRC2, authors aged 50-59 years comprised the highest proportion of first authors (36%) while in BRC3 authors aged 40-49 years made up the highest proportion (33%). In the last authors category, 55% was aged 40-49 years in BRC2 and 48% was aged 50-59 years in BRC3. The proportion of first authors aged 39 and younger increased in BRC3 compared to BRC2 while their proportion as last author decreased in BRC3 than in BRC2. For the total study period (2012-2022), there were statistically significant differences in the proportions of first and last authors by age of authors (p<0.001), showing the highest proportions of first and last authors in the age groups 40-49 and 50-59 years respectively (Table 1).

Percentage change in authorship positions in BRC3 compared to BRC2 (Figure 2) showed a substantial increase in first authorship by ethnicity, sex and age of authors except for authors aged 65 years and above. In BRC3, there was also a notable increase in last authorship by sex, ethnicity and age for all authors except for authors under 39 years of age who decreased by 2.4% compared to BRC2 (Figure 2).

**Figure 2.**
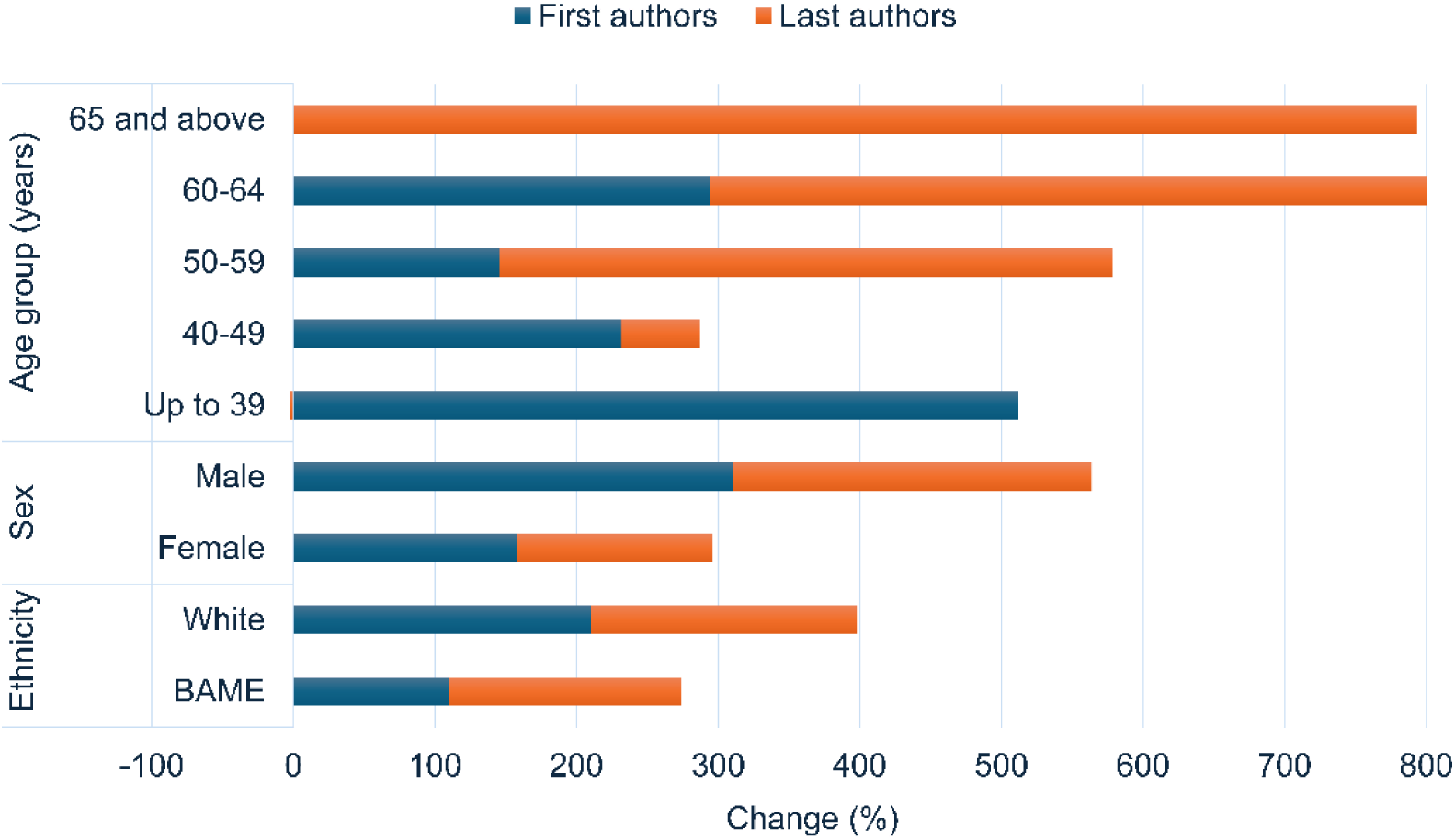
Percentage change in authorship positions in BRC3 compared to BRC2

### Publications by ethnicity, sex and age of authors

Publication output by author protected characteristics is presented in Table 2, which also shows disparities between authors of different ethnic backgrounds, genders and age groups. Publications by white authors were about nine times higher than publications by BAME authors in both BRCs, and the overall proportions of publications by author ethnicity were the same (white authors 87% and BAME authors 13%) in BRC2 and BRC3 (p=0.91). Male authors’ publications were statistically significantly more than female authors’ publications in both BRCs (p<0.001). Proportions of publications by authors of different age groups were statistically significantly different in both BRCs (p**<**0.001). Publications by authors aged 50-59 years were more than publications by authors of any other age group in both BRCs. Total publications substantially increased irrespective of ethnicity, sex and age of authors in BRC3 compared to BRC2 (Table 2). In the total study period (2012-2022), authors who were of white ethnicity, male gender and 50-59 years age produced the highest proportions of publications than authors of any other protected characteristic included in the study (Table 2).

**Table 2.** Publications and citations by ethnicity, sex and age of authors.

|  | Publications |  |  |  | Citations |  |  |  |
| --- | --- | --- | --- | --- | --- | --- | --- | --- |
| Author characteristics | BRC2<br>(2012–2017) | BRC3<br>(2017-2022) |  | Total study period<br>(2012-2022) | BRC2<br>(2012–2017) | BRC3<br>(2017-2022) |  | Total study period<br>(2012-2022) |
|  | Count (%) | Count (%) | P-value | Count (%) | Count (%) | Count (%) | P-value | Count (%) |
| **Ethnicity** |  |  |  |  |  |  |  |  |
| BAME | 263 (13) | 723 (13) | 0.91† | 986 (13) | 18439 (11) | 26761 (13) | <0.001† | 45200 (12) |
| White | 1777 (87) | 4835 (87) |  | 6612 (87) | 149341 (89) | 172951 (87) |  | 322292 (88) |
| **Sex** |  |  |  |  |  |  |  |  |
| Female | 891 (41) | 1792 (27) | <0.001† | 2683 (31) | 62217 (35) | 75516 (32) | <0.001† | 137733 (34) |
| Male | 1285 (59) | 4816 (73) |  | 6101 (69) | 113646 (65) | 158948 (68) |  | 272594 (66) |
| **Age group (years)** |  |  |  |  |  |  |  |  |
| Up to 39 | 72 (3) | 461 (7) | <0.001† | 533 (6) | 5541 (2) | 13595 (6) | <0.001† | 19136 (3) |
| 40-49 | 399 (18) | 1736 (27) |  | 2135 (25) | 55122 (17) | 73590 (33) |  | 128712 (23) |
| 50-59 | 1005 (46) | 2731 (42) |  | 3736 (43) | 129299 (39) | 87097 (38) |  | 216396 (39) |
| 60-64 | 436 (20) | 938 (15) |  | 1374 (16) | 97050 (29) | 40568 (18) |  | 137618 (25) |
| 65 and above | 263 (12) | 564 (9) |  | 827 (10) | 43578 (13) | 11112 (5) |  | 54690 (10) |
| †Pearson's Chi-squared test of Independence; BAME = Black, Asian and Minority Ethnic; BRC = Biomedical Research Centre |  |  |  |  |  |  |  |  |

Percentage change in publications by ethnicity, sex and age of authors in BRC3 compared to BRC2 is shown in Figure 3. The percentage change in BRC3 publications by BAME authors (175%) was greater than change in white authors’ publications (172%). The change in publications by male authors (275%) was greater than change in female authors’ publications (101%). The change in publications by young authors aged up to 39 years (540%) was the highest among authors of all age groups included in the study (Figure 3).

**Figure 3.**
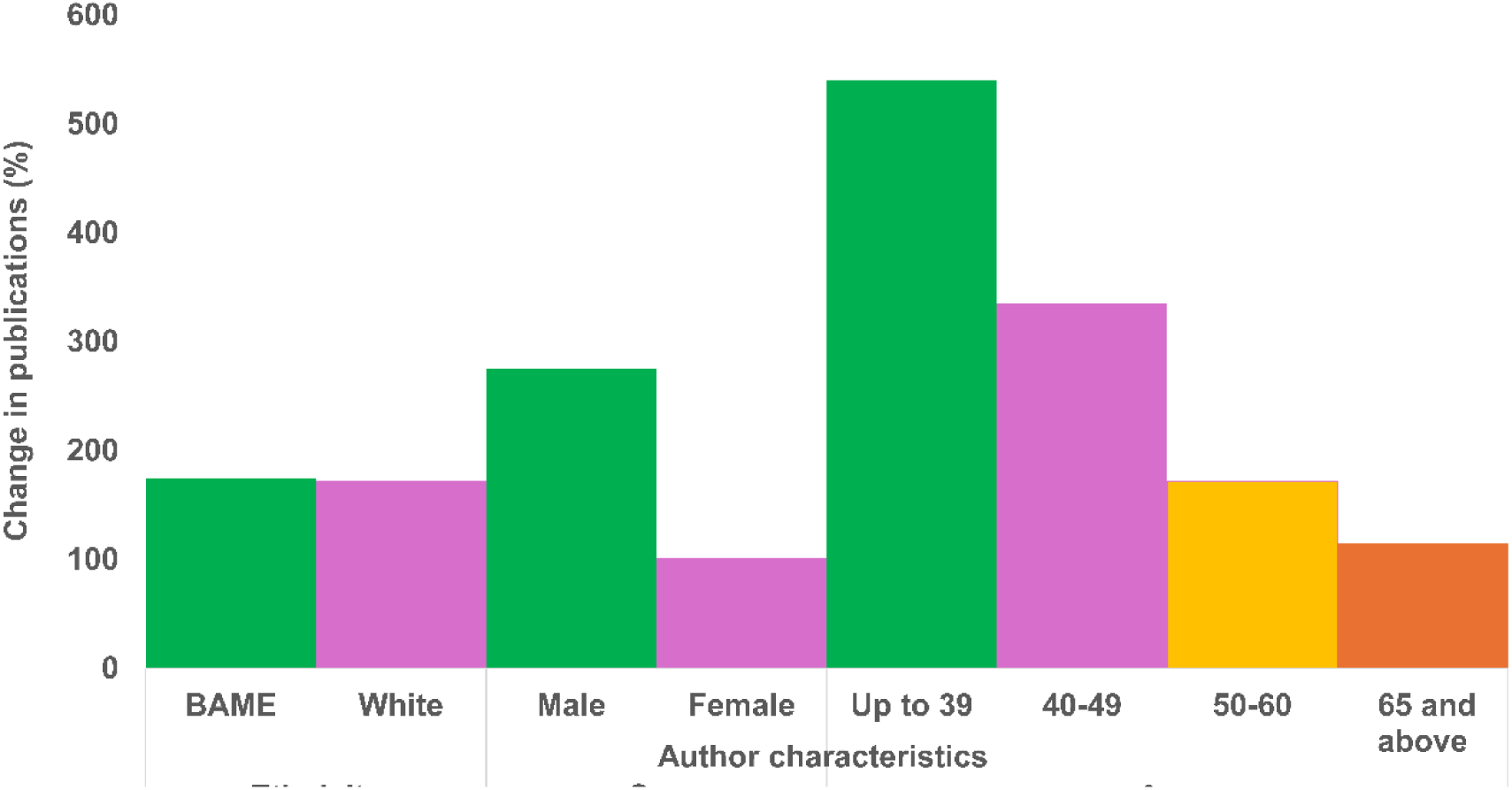
Percentage change in publications by ethnicity, sex and age of authors in BRC3 compared to BRC2

### Citations of publications by ethnicity, sex and age of authors

Citations of publications by ethnicity, sex and age of authors were statistically significantly different in BRC2 and BRC3 (p< 0.001), as shown in Table 2. The highest citations were for publications by authors who were of white ethnicity, male gender and 50-59 years age group in BRC2, BRC3 and total study period (Table 2). The absolute number of citations of BRC3 publications were higher than citations of BRC2 publications by authors of white and BAME ethnicities, male and female genders and all age groups except authors aged 50 years and above (Table 2).

Percentage change in total citations in BRC3 compared to BRC2 (Figure 4) showed that citations of publications by BAME authors increased (45%) more than citations of publications by white authors (16%) and citations of publications by male authors increased (40%) more than citations of publications by female authors (21%). Total citations of publications by authors aged up to 39 years and aged 40-49 years increased by 145% and 33% respectively; however, citations of publications by authors aged 50-59 years, 60-64 years, and 65 years and above decreased by 33%, 58% and 75% respectively in BRC3 compared to BRC2 (Figure 4).

**Figure 4.**
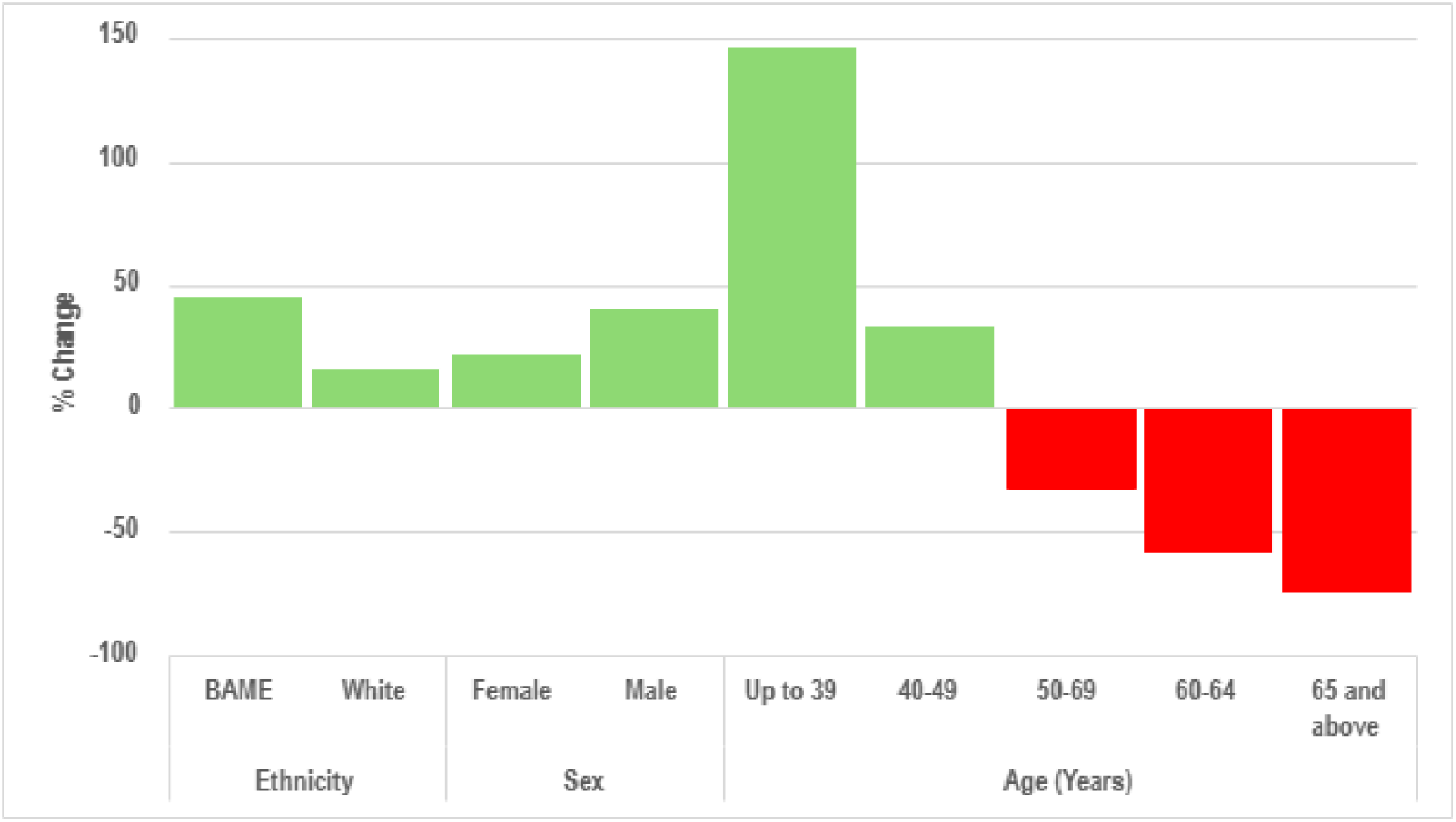
Percentage change in total citations in BRC3 compared to BRC2

Analysis of average citations per publication in BRC2 showed that authors who were white, male and aged 60-64 years had the highest citations compared to authors of other ethnicity, sex and age (Figure 5). In BRC3, female authors had higher citations per publication than male authors (42 versus 33 citations respectively) (Figure 5). In the total study period (2012-2022), citations per publication were not very different by ethnicity but female authors had higher citations than male authors and authors aged 60-64 years had the highest citations compared to authors of any other age group (Figure 5).

**Figure 5.**
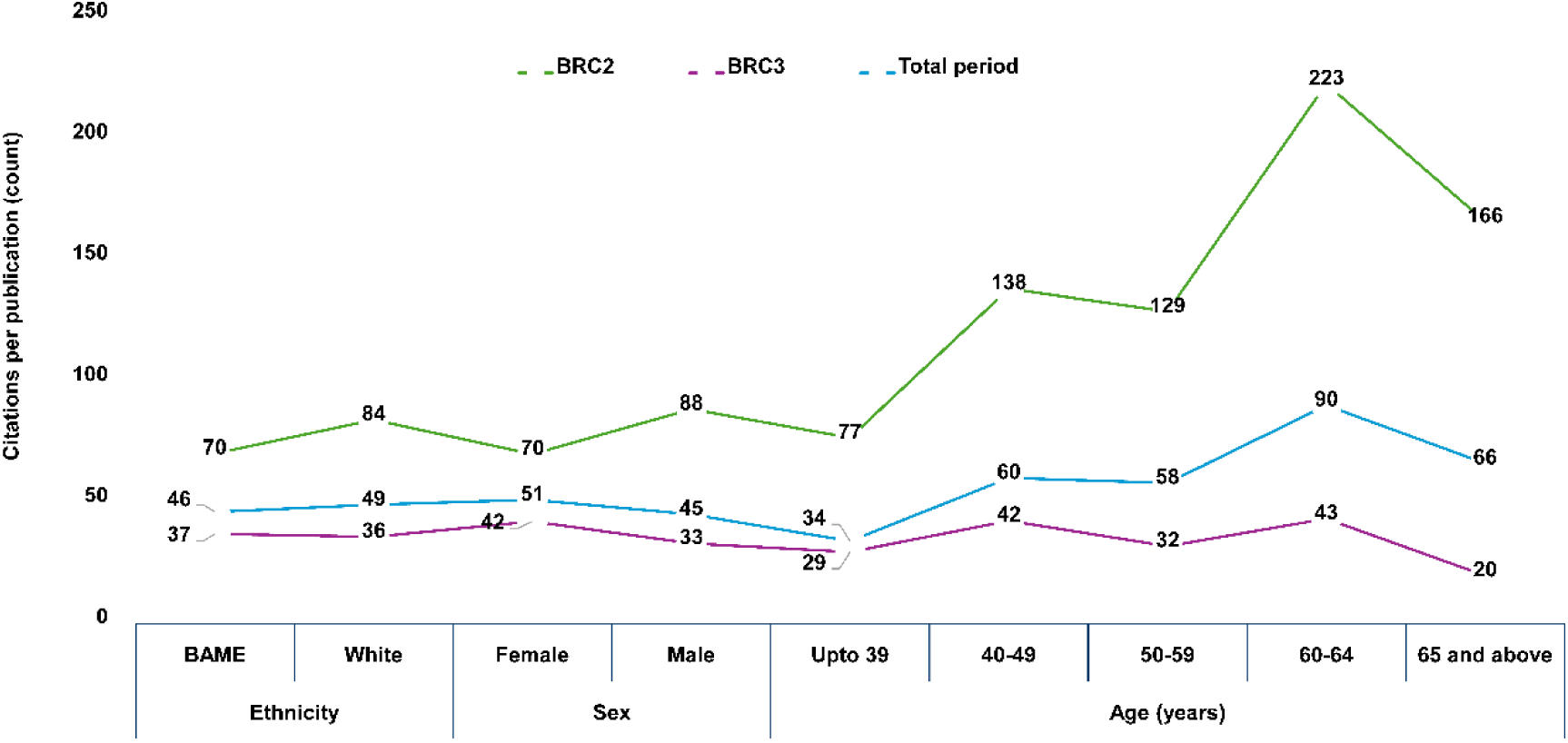
Average citations per publication by ethnicity, sex and age of authors

## DISCUSSION

We retrospectively analysed biomedical and translational health research publications funded by the NIHR Oxford BRC between April 2012 and November 2022. We analysed research publications at an individual researcher level, as it is the most common unit of analysis (17,18). We did this study at the environment level i.e., at the level of our BRC providing research facilities and funds (19), which are crucial factors in promoting research productivity (20,21) and fostering research collaboration (22). Our BRC is a large research infrastructure that supports innovative and translational health research. Latest evidence shows that such research support establishments generate high quality scientific research, innovation and collaborations (23).

We investigated three research publication metrics: (a) authorship position (first and last authors) as an indicator of research effort and prestige (24,25), (b) research publications as a measure of research productivity (26,27) and (c) publication citations as an indicator of the quality and impact of research in different disciplines (6,7), especially in Science, Technology, Engineering, and Mathematics (STEM) subjects including medicine (8). We analysed these publication metrics against three protected characteristics i.e., ethnicity, sex and age of authors supported by our BRC. Doing this study was imperative as there is limited understanding of and evidence on the diversity of authors of scientific research (28), especially in biomedical and translational health research (29). In addition, this study is important as it provides evidence on methodological challenges in research on diversity of scientific authorship (30).

### Author ethnicity

Our analysis of authorship type by author ethnicity showed that in both first and last authorship categories proportions of white authors were statistically significantly higher than the proportions of BAME authors in both BRC2 and BRC3 funding rounds and the total study period (Table 2). These findings are in corroborations with other studies that show white authors dominating in first and last authorships categories in western and developed countries like the US and the UK (31). The overall proportions of white first (84%) and last authors (86%) in the total period of our study (2012-2022) are slightly different from the proportions of white first (87.2%) and last authors (82.8%) found in 2012-2022 by Chander et al, (31).

Our findings show positive trends in the absolute number of BAME authors in both authorship categories in BRC3 compared to BRC2, but the proportions were not statistically significantly different. Proportions of BAME first (16%) and last authors (14%) in the total period (2012-2022) of our study were slightly lower than proportions of non-white authors reported by others (31). However, these differences could be country and institution specific as our findings are related to the NIHR Oxford BRC in England while findings reported by Chander et al. (31) relate to different institutions in different countries including the UK.

Findings of our analysis of number of publications by author ethnicity showed that nearly nine out of ten publications were published by white authors, which was due to the greater number of white researchers in both BRC funding periods. However, our findings of percentage change in number of publications by authors ethnicity in BRC3 compared to BRC2 revealed 175% increase in publications by BAME authors compared to 172% increase in publications by white authors, but these differences were not statistically significant (Table 2, Figure 3).

Our analysis of citations per publication by author ethnicity (Table 2) showed that in BRC2 period publications by white authors received higher citations (80 citations per publication) compared to publications by BAME authors (70 citations per publication). However, in BRC3 period, publications by BAME authors were cited slightly more than publications by white authors (37 vs 36 citations per publication respectively) (Figure 5). The difference in citations of publications by white and BAME authors could be due to biases in citations, which occur due to several factors such as the gender, ethnicity/race and country / nationality of the author (32–34).

### Author gender

Our analysis of publication metrics by author sex (male and female only) showed statistically significant differences between male and female authors in first and last authorship positions. These findings are in conformity with earlier studies (9,35,36). We found a greater number of male authors as first and last authors in both BRC2 and BRC3. These findings are similar to several other studies that show higher proportions of male authors in prestigious authorship positions in medical and translational health research (9,29). However, we found some positive trends showing increase in the absolute number of female authors as first and last authors in BRC3 compared to BRC2; however, it was lower than the increase in male first and last authors in BRC3 (Table 1, Figure 2). Similar positive trends in gender equity are also reported by others (36); but the gender gap in authorships remains to be narrowed down (37–39).

Our findings of publication count by author sex (Table 1) showed that total number of publications by male authors were statistically significantly greater than publications by female authors in both BRC funding rounds, as reported in earlier studies (9). We noted a positive trend showing increase in the publications output by female authors in BRC3 compared to BRC2 (Table 2, Figure 3); but the increase was lower than the proportional increase in publications by male authors. Our findings corroborate earlier studies reporting the underrepresentation of female authorship in biomedical publications in the UK (9,13). Our earlier study on the gender equity in the authorship of scientific publications found the overall proportion of female authors lower than male authors but also significant increasing trends of female first, last and corresponding authors while the proportions of male and female last authors were similar to their respective proportions as principal investigators in our BRC (9). A recent Elsevier gender report (12) reported that on average, women researchers publish fewer publications than men in every country participating in the study, regardless of authorship position. They reported the least difference in the number of publications by women compared to men among first authors while the biggest difference were observed among all other authorships categories (12).

Despite evidence of the benefits of diversity both for the research organisations and the research quality, our findings illustrate and confirm that there are profound author diversity biases in both publication and citation practices (40,41). These biases suggest lack of EDI at the author level (42). There is evidence of demographic haemophilia and relations between the editors, reviewers and the authors (43,44), especially in the biosciences (45). This implies that editors and reviewers prefer authors of the same gender and nationality (44,45), which may provide part of the explanation of our findings. In addition, earlier research concludes that overall male researchers are more productive than female researchers but the gender differences in publications are affected by a combination of factors such as the age and position of the researcher, access to funding and obtaining prestigious research grants, opportunities to become independent researchers, and research environment (46). These factors could negatively impact researchers who are female, younger in age and from ethnic minority backgrounds, as our findings show in this study.

Part of the explanation of our findings as to the research productivity differences due to author gender may be the fact that male researchers, especially last authors are older in chronical age, have longer academic age and occupy higher academic positions (47,48). These factors are associated with many advantages such as access to research funding, research networks and collaboration, and have high impact research profile and publication portfolio (49). Conversely, there are few female scientists at higher positions and they are often less in prestigious authorship positions compared to male researchers (1,9). There are also widening gender disparities in research projects (50,51), clinical-trial leadership (52) and innovation and creativity as evident by global patenting trends showing women comprising only 23% of all applicants (38). These factors have a negative impact on women scientists’ research performance (53), scientific publication productivity (46) and research impact (54,55). Women scientists need guidance, support and mentoring, and research funding opportunities to excel in scientific research and innovation (56).

We also found statistically significant differences in citations of publications by author sex showing overall higher proportions of citations of publications by male authors than female authors in each BRC period (Table 2). This is not surprising as similar trends showing a gender citation gap are reported by others (25,57). However, our analysis of the cumulative citations per publications over the total period of our study (2012–2022) revealed that the average citations per publication by female authors were 51 citations compared to 45 citations per publication by male authors (Figure 5). In addition, in BRC3 period, publications by female authors were cited more (42 citations per publications) than publications by male authors (33 citations per publication) (Figure 5). These findings indicate a positive trend, which may also corroborate other studies that suggest that female scientists publish less but could get higher citations (58), indicating women scientists’ excellence in research and innovation (59).

Our findings also confirm differences in the research publications productivity and citations of publications by author gender (female and male). Our study show that women authors have fewer publications but more citations compared to men authors, as reported by others (58); while other studies show gender bias in citation practice, particularly among first authors showing higher average citation impact of men compared to women (12). These findings contribute to the body of research that shows citation inequality in medical sciences (60,61) and a greater percentage of citations accumulated by a small group of elite scientists (61); thus, having the Matthew effect in their research (62,63). However, variations in citations by author gender are small in medicine literature. The citation picture is more complex and the association between research productivity, quality and impact of research is not specific to author gender only (46). There are several contributing factors such as unethical citation practices such as self-citation, journal prestige (60,64), research policies and cultural norms (65).

### Author age

Study of association between author age and publication metrics has been focus of several bibliometric studies (49,66) such as author age and authorship positions (67), author age and publication productivity (48,68), author age and citations (48,66), as well as EDI in scientific authorship (69). We studied author age and trends in three publication metrics i.e., authorship positions, publication output and citations. We used the chronological age of authors; whereas most studied have used academic age as a proxy of author age (47,48). Out findings showed statistically significant differences between authors of different age groups and three publication metrics. We found statistically significant differences in first and last authorship positions by author age in BRC2 and BRC3 (Table 2). For the total period of our study (2012-2022), we found a substantial proportion of first authors under the age of 40 years, the proportion increased in 40s and then decreased in 50s onwards. Conversely, in the last author category, we found a very small proportion of young authors (under 40 years of age) while the proportion of last authors increased from the age of 40 years, reached at the peak in 50s and then decreased in 60s onwards (Table 2; Figure 6 – panel A). These findings suggest that the relationship between authors age and first and last authorship positions is quadratic and non-linear (67,69).

**Figure 6.**
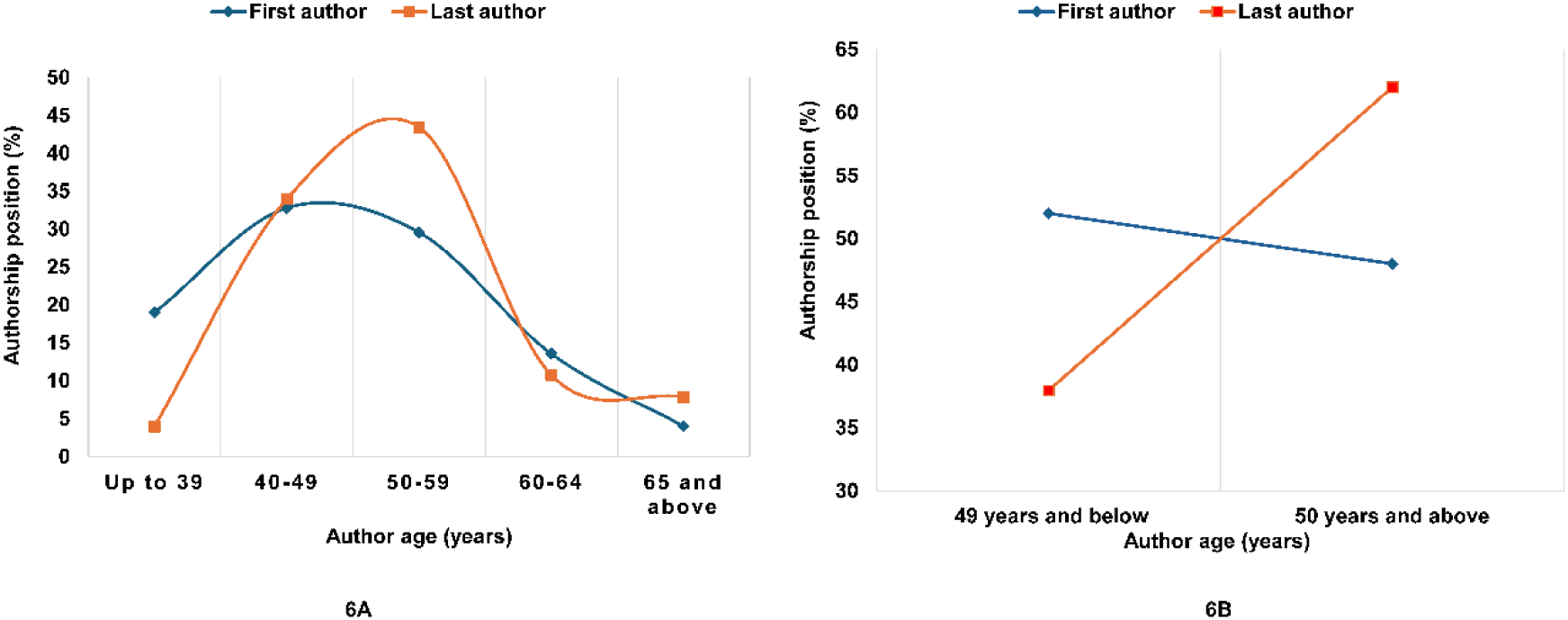
First and last authorship positions by author age (Total study period 2012-2022)

Interestingly, when we divided first and last authors into two age groups: (a) 49 years and below, and (b) 50 years and above, we found a clear divide in first and last positions by author age; however, the difference was not statistically significant (p=0.064). Authors aged 49 years and below comprised the highest proportion of first authors and the lowest proportion of last authors while authors aged 50 years and above comprised the highest proportion of last authors and lowest proportion of first authors (Figure 6-panel B). These findings are in conformity with earlier research that first authors are more likely to be young researchers while the last authors are highly likely to be old researchers (48,69). However, the relationship between author age and authorship positions is complex because the authorship position is determined and influenced by several factors. For example, the author’s academic age, experience, rank and role as well as other factors like research team and network, academic discipline, institutional policies and the authorship order culture in academic disciplines and research institutions (47,70–72).

Our findings also show statistically significant differences in publication output by authors of different age groups in BRC2, BRC3 and total study duration. We found that authors aged 50-59 years and 40-49 years were the first and second most productive authors, who published the first and second highest proportions of publications respectively in BRC2 and BRC3 (Table 2). These findings are not surprising as earlier research shows that middle-aged authors are most likely to publish higher numbers of publications than authors of any other age (48). Because of the higher publication productivity, author age of 40-59 years has been called as the golden age of authors (73) and our findings provide evidence that authors of this age group produced 68% of total publications (64% in BRC2 and 69% in BRC3) in our BRC during the study period (Table 2). We also found that authors aged under 40 years produced the smallest proportion of publications, as reported by others (48,68,69). Young early career researchers are more likely to produce a smaller number of publications because they face several challenges. For example, short-term research contracts, lack of research training, mentorship and infrastructural support, often work alone on research projects with limited protected time, and encounter barriers in research publication and funding (74).

Contrary to earlier studies suggestions that publication productivity of authors does not decline in older age (48,75), our findings show declining trends in publication output by older authors aged 60 and above, as reported by others (69). The declining trend in publication productivity of older authors could be due to various reasons such as a higher pressure to acquire funding and less pressure to write research articles (76) and a change in the nature of research activity and focus to some other scholarly work (75). Overall, our findings show an inverted U shape like concave relationship between the author age and research publications (77), which suggests that publication productivity increases with author age, plateaus in 50s and then declines in 60s and onwards (48,69,78). However, this type of relationship between author age and publication productivity has been contradicted by some studies, which suggest that this pattern applies to some researchers but does not hold universally (68,79).

The relationship between author age and publication citations is also complex. Our findings show a curvilinear relationship between the author age and total citations. We found that authors aged 50-59 years had the highest proportion of total citations than authors of any other age group in BRC2, BRC3 and the total period of our study (Table 2). It is not surprising because the age of publication usually determines the number of citations; thus, older publications have higher citations than the newer publications (66). Very interestingly, we found that citations of BRC3 publications of younger authors (less than 50 years of age) increased by manifold while older authors’ citations decreased despite having the same publication age (BRC3, 2017-2022) (Figure 3). This positive trend in higher citations of younger authors’ recent publications in BRC3 period could be due to various factors (80), mainly related to the author, journal and publication (81). For example, the content of publication; high importance, innovation and growth of the topic; impact of research and accessibility e.g. open access of publications; and the structural context of research like publications from large prestigious research institutions (82,83).

More interestingly, we also found that authors aged 60-64 years had the highest average citations per publication in both BRCs as well as in the total study period (Figure 5). These findings suggest that older authors are usually senior and reputed researchers and well known in areas of their research specialism; they usually lead research teams, often have (inter-) national research collaborations and networks; their publications have higher visibility and accessibility; hence, they get quick attention and higher citations (84–86).

We also found that authors aged 60 to 64 years had averagely higher citations of their publications of the same age compared to authors of any other age group and their citations were averagely 20 citations per publication higher than average citations per publication by authors of any other age. The reason could that authors of this age (60 to 64 years) are usually senior researchers and have highly advantageous positions. They might be consultants, research group leaders, leading several projects. They might be well known authorities locally, national and if not globally. Therefore, their publications from their local research group, and/or from a national or an international collaboration are highly likely to get quick and higher visibility soon after publications. Most of the authors of this age usually work on cutting age techniques and procedures in biomedical and translational health research domain, which is the focus of our BRC funded research and our BRC is one of the world-famous centres for translational and biomedical research.

Our study demonstrates significant disparities in authors positions, research publications and citations by author ethnicity, sex and age. Our findings align with recent research suggesting widening author-level disparities in medical sciences (37,61). These disparities are determined by a variety of complex factors such as the research environment, funding frameworks, research collaborations and networks, and fields/disciplines characteristics (87).

Our findings should not be looked at in isolation but need to be judged keeping in view the broader context at the national, sectoral, organisational, higher education, research funding and disciplinary levels. At the national level, the ethnic profile of the population in England shows that white ethnic population comprises the highest proportion of the total population (85% and 81% in 2011 and 2021 censuses, respectively) and the ethnic minorities comprise a smaller proportion of the population (88). Our findings also show a similar picture as most of the first and last authors were of white ethnicity (Table 1). In addition, the diversity in higher education staff data shows that a vast majority of professors are male (68% vs 32% female) and white (86%) (white male 53% and white female 26%) compared to 14% professors of ethnic minority backgrounds (89). Moreover, employment in professional occupations in STEM disciplines are dominated by male professionals (75% vs 25% women (90). These inequalities in the higher education sector and professional occupations in STEM fields are reflected in our findings about the gender and ethnicity of authors. Furthermore, in the healthcare sector, we find 69% white clinicians and 32% of Asian clinicians (43.5% associate specialists and 30% consultants) (91). However, these proportions of clinicians of ethnic minority backgrounds are not depicted in the authorship positions in our study. The other sector that we should look at is the employment of researchers. Our BRC is a non-recruiting research centre, which is a partnership between the University of Oxford and Oxford University Hospitals NHS Foundation Trust. Hence researchers funded by our BRC are employees of either the University or the hospital trust. Staff diversity data of the University of Oxford shows more male academics (65%) and researchers (53%) than their female counterparts (35% academics and 47% researchers) and about 68% of academics and 54% of researchers are white compared to BAME academics (11%) and researchers (27%) (92). A similar picture of staff diversity is present at the Oxford University Hospital NHS Trusts where clinical staff is 70% white and about 30% BAME staff (93). Our findings showing disparities in publication metrics by author ethnicity, gender and age have roots in the above-mentioned broader contexts. In addition, we must also consider the broader funding mechanisms. Our BRC is funded by the NIHR, and their award funding data shows that only 12% applicants of ethnic minority backgrounds get research funding compared to 88% white applicants, and 64% fundings is awarded to researchers aged 40 and 59 years while the proportion of research awards by gender are similar (61% female and 59% male) (94). This means that applicants who are younger in age and belong to ethnic minority backgrounds are less likely to secure research funding necessary to pursue their research pathways. All this evidence suggests that there is a need for proactive initiatives to facilitate, support, promote and provide fairer and equitable opportunities to disadvantaged researchers who are young, female and belong to ethnic minority backgrounds (19,95). Tackling these inequalities requires reinforcing institutional commitments to EDI in research at recruitment, training, career progression, authorship and funding levels through policies, programmes, training, retention and promotion of researchers of diverse ethnic backgrounds, genders and age groups (96–98). Such actions would require appropriate and proactive efforts at different levels including employers and research organisations and research funders (99). This would also need strategic initiatives and policies such as the EDI strategies in research institutions and funding organisations. Some positive initiatives have been taken such as the formulation of EDI strategies by our BRC (100) and Oxford University NHS Hospitals NHS Foundation Trust (101) and the EDI strategic plan by the University of Oxford (102). Our BRC funder, the NIHR also has an EDI strategy (3). However, these strategies would take some time to narrow down the disparities in research. However, for tracking progress on EDI, it would be imperative to collect author / researcher level diversity data including data on protected characteristics at the time of recruitment as well as data on publication metrics for reporting to the research funder. In this way a fairer equality, diversity and inclusion reflecting the actual situation in the sector specially in medicine and health research would be facilitated and realised.

### Study Limitations

This study is not without limitations and challenges. The main limitations include the use of self-reported data on protected characteristics provided by researchers during the recruitment process and collated by the HR Department of the University of Oxford. These data were not specifically collected for tracking diversity of authors (researchers) funded by our BRC. Data on protected characteristics was provided to us in an aggregated form by the University HR Department and there was no data on protected characteristics for some authors. This is because reporting data on protected characteristics is not mandatory but voluntary (1) and acquiring and collating such data is difficult and complicated (5). Therefore, some authors could have been excluded from the analysis. Nonetheless, we analysed a large dataset that included three protected characteristics of BRC funded authors (476 unique researchers) and three important publication metrics covering about a decade (2012 to 2022).

## CONCLUSION

Our study provides important evidence on diversity of authors in biomedical and translational health research funded by the NIHR Oxford BRC. Our findings confirm inequality in authorship, publication productivity and citations, three important metrics of biomedical and translational health research at the level of author ethnicity gender, and age. In order to achieve equality, diversity and inclusion at an NHIR BRC level, it is imperative to address structural inequalities (3,103) through systematic investigation of factors such as recruitment, retention, training, career progression and promotion and research funding barriers (5,104), which have a great potential to contribute to inequalities and exclusions of researchers at different levels in the NIHR BRCs (105).

In this study, we were reliant on researchers’ self-reported protected characteristics data provided as part of the recruitment process and collated by the HR Department. Improved collation of such data would provide important evidence to determine the denominator of researchers with such protected characteristics being recruited and facilitate clearer actions to address potential recruitment bias of researchers with such protected characteristics. In addition, collecting diversity data by ethnicity, age and sex of authors of publications funded by NIHR BRCs would be helpful in tracking progress on EDI in biomedical and translational research. Further research is required to improve data quality and ways of capturing diversity of research authorship.

## Author contributions

LRH conceived the study, drafted the manuscript, reviewed and implemented feedback from all other authors.

FEB and DS extracted publication data matched with authors from the Symplectic Elements current research information system (CRIS) used by the University of Oxford

OC supported in collating peer-reviewed publications of researchers funded by the NIHR Oxford BRC.

VK supported the study and arranged funding for the study by the NIHR Oxford BRC.

EKS conceived the study, reviewed the manuscript and provided critical input.

SGSS conceived the study, analysed data, created visualisations and drafted, edited and finalised the manuscript.

## Disclaimer

The views expressed are those of the authors and not necessarily those of the NHS, the NIHR or the Department of Health and Social Care.

## Ethics approval

The University of Oxford Clinical Trials and Research Governance Team reviewed the study and deemed it exempt from full ethics review on the grounds that it is deemed a service development project (date: 16 September 2021).

## Data Availability

Data on publications and citations are available from authors upon reasonable request. However, due to the protected nature, authors’ protected characteristics data is not available.

## REFERENCES

1. Henderson LR, Dam R, Shah SGS, Ovseiko PV, Kiparoglou V. Perceptions of gender equity and markers of achievement in a National Institute for Health Research Biomedical Research Centre: a qualitative study. Health Res Policy Syst [Internet]. 2022 Sept 24;20(1):102. Available from: 10.1186/s12961-022-00904-4

2. National Institute for Health and Care Research (NIHR). Diversity Data Report 2022 [Internet]. London: Department of Health and Social Care; 2024 Sept. Available from: https://www.nihr.ac.uk/about-us/who-we-are/research-inclusion/diversity-data

3. National Institute for Health and Care Research (NIHR). Equality, Diversity and Inclusion Strategy 2022-2027 [Internet]. London: Department of Health and Social Care; 2022 Sept. Available from: https://moorfieldsbrc.nihr.ac.uk/case-study/edi-strategy-2022-2027/

4. Rice DB, Raffoul H, Ioannidis JPA, Moher D. Academic criteria for promotion and tenure in biomedical sciences faculties: cross sectional analysis of international sample of universities. BMJ [Internet]. 2020 June 25;369:m2081. Available from: http://www.bmj.com/content/369/bmj.m2081.abstract

5. Henderson LR, Shah SGS, Ovseiko PV, Dam R, Buchan AM, McShane H, et al. Markers of achievement for assessing and monitoring gender equity in a UK National Institute for Health Research Biomedical Research Centre: A two-factor model. PLoS One [Internet]. 2020 Oct 14;15(10):e0239589. Available from: 10.1371/journal.pone.0239589

6. Leydesdorff L, Bornmann L, Comins JA, Milojević S. Citations: Indicators of Quality? The Impact Fallacy. Front Res Metr Anal [Internet]. 2016;1:1. Available from: 10.3389/frma.2016.00001

7. Aksnes DW, Langfeldt L, Wouters P. Citations, Citation Indicators, and Research Quality: An Overview of Basic Concepts and Theories. SAGE Open [Internet]. 2019 Jan 1 [cited 2025 Apr 23];9(1):2158244019829575. Available from: 10.1177/2158244019829575

8. Thelwall M, Kousha K, Stuart E, Makita M, Abdoli M, Wilson P, et al. In which fields are citations indicators of research quality? J Assoc Inf Sci Tech [Internet]. 2023 Aug 1 [cited 2025 Nov 28];74(8):941–53. Available from: 10.1002/asi.24767

9. Shah SGS, Dam R, Milano MJ, Edmunds LD, Henderson LR, Hartley CR, et al. Gender parity in scientific authorship in a National Institute for Health Research Biomedical Research Centre: a bibliometric analysis. BMJ Open [Internet]. 2021 Mar 1;11(3):e037935. Available from: 10.1136/bmjopen-2020-037935

10. American Journal Experts (AJE). Durham, NC., USA. 2023. First Author vs. Corresponding Author? How to Decide Which to Choose. Available from: https://www.aje.com/arc/first-author-vs-corresponding-author/

11. Alfred J. Assigning Authorships [Internet]. Brighton, England: UK Research Integrity Office; 2023. Available from: https://ukrio.org/wp-content/uploads/Case-study-14-for-trainers_STEM-case-study-on-assigning-authorship.pdf

12. Van der Linden N, Browning E, Roberge G. Gender Equality in Research & Innovation-2024 Review [Internet]. Elsevier Data Repository; 2024. Available from: 10.17632/bb5jb7t2zv.2

13. Abdalla S, Abdalla M, Saad M, Jones D, Podolsky S, Abdalla M. Ethnicity and gender trends of UK authors in The British Medical Journal and the Lancet over the past two decades: a comprehensive longitudinal analysis. eClinicalMedicine [Internet]. 2023 Oct 1 [cited 2025 June 16];64. Available from: 10.1016/j.eclinm.2023.102174

14. Kang-Auger S, Ukah UV, Healy-Profitós J, Ayoub A, Auger N. Persistent ethnic disparities in authorship within top European and North American medical journals: a serial cross-sectional analysis. J Clin Epidemiol [Internet]. 2024 Dec 1;176:111552. Available from: 10.1016/j.jclinepi.2024.111552

15. NIHR Oxford Biomedical Research Centre. About the NIHR Oxford Biomedical Research Centre [Internet]. 2025. Available from: https://oxfordbrc.nihr.ac.uk/about-us-intro/

16. National Institute for Health and Care Research (NIHR). Biomedical Research Centres [Internet]. 2025. Available from: https://www.nihr.ac.uk/about-us/what-we-do/infrastructure/biomedical-research-centres

17. Sahel JA. Quality Versus Quantity: Assessing Individual Research Performance. Sci Transl Med [Internet]. 2011 May 25 [cited 2025 Apr 24];3(84):84cm13–84cm13. Available from: 10.1126/scitranslmed.3002249

18. Kwiek M, Roszka W. Are Scientists Changing their Research Productivity Classes When They Move Up the Academic Ladder? Innov High Educ [Internet]. 2025 Feb 1;50(1):329–67. Available from: 10.1007/s10755-024-09735-3

19. Kiparoglou V, Brown LA, McShane H, Channon KM, Shah SGS. A large National Institute for Health Research (NIHR) Biomedical Research Centre facilitates impactful cross-disciplinary and collaborative translational research publications and research collaboration networks: a bibliometric evaluation study. J Transl Med [Internet]. 2021 Nov 27;19(1):483. Available from: 10.1186/s12967-021-03149-x

20. Ocampo L, Aro JL, Evangelista SS, Maturan F, Yamagishi K, Mamhot D, et al. Research Productivity for Augmenting the Innovation Potential of Higher Education Institutions: An Interpretive Structural Modeling Approach and MICMAC Analysis. J Open Innov Technol Mark Complex [Internet]. 2022 Sept 1;8(3):148. Available from: https://www.sciencedirect.com/science/article/pii/S2199853122007491

21. Jürgens A, Tedeschi G, D’Errico G, Kilian K, Zawadzki K, Daniel O, et al. Navigating the frontier: research infrastructures, core facilities and a new paradigm at European Universities. Cogent Educ [Internet]. 2024 Dec 31;11(1):2365613. Available from: 10.1080/2331186X.2024.2365613

22. Abramo G, D’Angelo CA, Di Costa F. Research collaboration and productivity: is there correlation? High Educ [Internet]. 2009 Feb 1;57(2):155–71. Available from: 10.1007/s10734-008-9139-z

23. Xing Y, Wu Y, Xiao X, Wang D, Zhang LL. Mega research infrastructure as a driver for high-quality development and innovation: Promoting scientific cooperation and interdisciplinarity. Proj Leadersh Soc [Internet]. 2025 Dec 1;6:100150. Available from: 10.1016/j.plas.2024.100150

24. Peidu C. Can authors’ position in the ascription be a measure of dominance? Scientometrics [Internet]. 2019 Dec 1;121(3):1527–47. Available from: 10.1007/s11192-019-03254-1

25. Bendels MHK, Müller R, Brueggmann D, Groneberg DA. Gender disparities in high-quality research revealed by Nature Index journals. PLoS One [Internet]. 2018 Jan 2;13(1):e0189136. Available from: 10.1371/journal.pone.0189136

26. Abramo G, D’Angelo CA. How do you define and measure research productivity? Scientometrics [Internet]. 2014 Nov 1;101(2):1129–44. Available from: 10.1007/s11192-014-1269-8

27. Carpenter CR, Cone DC, Sarli CC. Using Publication Metrics to Highlight Academic Productivity and Research Impact. Acad Emerg Med [Internet]. 2014 Oct 1 [cited 2025 June 16];21(10):1160–72. Available from: 10.1111/acem.12482

28. Loui M, Fiala SC. Inequities in Academic Publishing: Where Is the Evidence and What Can Be Done? Am J Public Health [Internet]. 2024 Apr 1 [cited 2025 June 16];114(4):377–81. Available from: 10.2105/AJPH.2024.307587

29. Charpignon ML, Matos J, Nakayama LF, Gallifant J, Alfonso PGI, Cobanaj M, et al. Diversity in the medical research ecosystem: a descriptive scientometric analysis of over 49 000 studies and 150 000 authors published in high-impact medical journals between 2007 and 2022. BMJ Open [Internet]. 2025 Jan 1;15(1):e086982. Available from: 10.1136/bmjopen-2024-086982

30. Sofi-Mahmudi A, Stojanova J, Vounzoulaki E, Tomlinson E, Pizarro AB, Khademioore S, et al. Cochrane reviews’ authorship has become more gender-diverse but remains geographically concentrated: a meta-research study. J Clin Epidemiol [Internet]. 2025 May 1;181:111719. Available from: 10.1016/j.jclinepi.2025.111719

31. Chander S, Luhana S, Sadarat F, Leys L, Parkash O, Kumari R. Gender and racial differences in first and senior authorship of high-impact critical care randomized controlled trial studies from 2000 to 2022. Ann Intensive Care [Internet]. 2023 June 27;13(1):56. Available from: 10.1186/s13613-023-01157-2

32. Ray KS, Zurn P, Dworkin JD, Bassett DS, Resnik DB. Citation bias, diversity, and ethics. Account Res [Internet]. 2024;31(2):158–72. Available from: 10.1080/08989621.2022.2111257

33. Kozlowski D, Murray DS, Bell A, Hulsey W, Larivière V, Monroe-White T, et al. Avoiding bias when inferring race using name-based approaches. PLoS One [Internet]. 2022 Mar 1;17(3):e0264270. Available from: 10.1371/journal.pone.0264270

34. Gomez CJ, Herman AC, Parigi P. Leading countries in global science increasingly receive more citations than other countries doing similar research. Nat Hum Behav [Internet]. 2022 July 1;6(7):919–29. Available from: 10.1038/s41562-022-01351-5

35. Chander S, Sorath F, Mohammed YN, Parkash O, Sadarat F, Lohana AC, et al. Gender, Race, and Regional Disparities in Leading Authorships of Gastroenterology and Hepatology Randomized Controlled Trials. J Gastrointest Cancer [Internet]. 2025;56(1):34. Available from: 10.1007/s12029-024-01161-0

36. Rampersad C. Female authorship trends in a high-impact Canadian medical journal: a 10-year cross-sectional series, 2013–2023. BMJ Open [Internet]. 2025 May 1;15(5):e093157. Available from: 10.1136/bmjopen-2024-093157

37. Madsen EB, Nielsen MW, Bjørnholm J, Jagsi R, Andersen JP. Author-level data confirm the widening gender gap in publishing rates during COVID-19. eLife [Internet]. 2022 Mar 16;11:e76559. Available from: 10.7554/eLife.76559

38. World Intellectual Property Organization (WIPO). The Global Gender Gap in Innovation and Creativity: An International Comparison of the Gender Gap in Global Patenting over Two Decades [Internet]. Geneva, Switzerland: Department for Economics and Data Analytics.; 2023. (WIPO Development Studies). Available from: https://www.wipo.int/publications/en/details.jsp?id=4653

39. Lauper K, Buitrago-Garcia D, Courvoisier DS, Iudici M, Mongin D. Trends and influences in women authorship in randomised controlled trials in rheumatology: a comprehensive analysis of all published RCTs from 2009 to 2023. RMD Open [Internet]. 2025 Mar 27;11(1):e005341. Available from: 10.1136/rmdopen-2024-005341

40. Gøtzsche PC. Citation bias: questionable research practice or scientific misconduct? J R Soc Med [Internet]. 2022 Jan 1 [cited 2025 June 16];115(1):31–5. Available from: 10.1177/01410768221075881

41. Urlings MJE, Duyx B, Swaen GMH, Bouter LM, Zeegers MP. Citation bias and other determinants of citation in biomedical research: findings from six citation networks. J Clin Epidemiol [Internet]. 2021 Apr 1;132:71–8. Available from: 10.1016/j.jclinepi.2020.11.019

42. Rouan J, Velazquez G, Freischlag J, Kibbe MR. Publication bias is the consequence of a lack of diversity, equity, and inclusion. J Vasc Surg [Internet]. 2021 Aug 1 [cited 2025 June 16];74(2):111S–117S. Available from: 10.1016/j.jvs.2021.03.049

43. Torre M, Prieto-Alonso JA, Ucar I. The uneven effects of gender parity: Trends in gender homophily in scientific publications, 1980–2019. Soc Sci Res [Internet]. 2025 Nov 1;132:103228. Available from: 10.1016/j.ssresearch.2025.103228

44. Kwiek M, Roszka W. Gender-based homophily in research: A large-scale study of man-woman collaboration. J Informetr [Internet]. 2021 Aug 1;15(3):101171. Available from: 10.1016/j.joi.2021.101171

45. Murray D, Siler K, Larivière V, Chan WM, Collings AM, Raymond J, et al. Author-Reviewer Homophily in Peer Review. bioRxiv [Internet]. 2019 Jan 1;400515. Available from: 10.1101/400515

46. van den Besselaar P, Sandström U. Vicious circles of gender bias, lower positions, and lower performance: Gender differences in scholarly productivity and impact. PLoS One [Internet]. 2017 Aug 25;12(8):e0183301. Available from: 10.1371/journal.pone.0183301

47. Chinchilla-Rodríguez Z, Escabias M, Morillo F. Meeting evaluation criteria based on authorship positions by gender, academic age, and research field: the Spanish case. Res Eval [Internet]. 2025 Jan 1 [cited 2025 Nov 23];34:rvaf008. Available from: 10.1093/reseval/rvaf008

48. Gingras Y, Larivière V, Macaluso B, Robitaille JP. The Effects of Aging on Researchers’ Publication and Citation Patterns. PLoS One [Internet]. 2009 Dec 29;3(12):e4048. Available from: 10.1371/journal.pone.0004048

49. Kwiek M, Roszka W. Academic vs. biological age in research on academic careers: a large-scale study with implications for scientifically developing systems. Scientometrics [Internet]. 2022 June 1;127(6):3543–75. Available from: 10.1007/s11192-022-04363-0

50. Liao CH, Lian JW. Gender inequality in applying research project and funding. J Inf Sci [Internet]. 2024 Apr 1;50(2):546–54. Available from: 10.1177/01655515221097861

51. Gao J, Yin Y, Myers KR, Lakhani KR, Wang D. Potentially long-lasting effects of the pandemic on scientists. Nat Commun [Internet]. 2021 Oct 26;12(1):6188. Available from: 10.1038/s41467-021-26428-z

52. Cevik M, Haque SA, Manne-Goehler J, Kuppalli K, Sax PE, Majumder MS, et al. Gender disparities in coronavirus disease 2019 clinical trial leadership. Clin Microbiol Infec [Internet]. 2021 July 1;27(7):1007–10. Available from: 10.1016/j.cmi.2020.12.025

53. Todeschini R, Baccini A. Handbook of bibliometric indicators: Quantitative tools for studying and evaluating research [Internet]. Weinheim: Wiley; 2016. Available from: 10.1002/9783527681969

54. Schneider M, Kane CM, Rainwater J, Guerrero L, Tong G, Desai SR, et al. Feasibility of common bibliometrics in evaluating translational science. J Clin Transl Sci [Internet]. 2017;1(1):45–52. Available from: 10.1017/cts.2016.8

55. Sandström U, van den Besselaar P. Quantity and/or Quality? The Importance of Publishing Many Papers. PLoS One [Internet]. 2016 Nov 21;11(11):e0166149. Available from: 10.1371/journal.pone.0166149

56. Heidt A. Behind every great woman in science, there’s another great woman in science. Nature [Internet]. 2025;639(8053):261–4. Available from: 10.1038/d41586-025-00620-3

57. Wu C. The gender citation gap: Approaches, explanations, and implications. Sociol Compass [Internet]. 2024 Feb 1 [cited 2025 June 16];18(2):e13189. Available from: 10.1111/soc4.13189

58. Campbell LG, Mehtani S, Dozier ME, Rinehart J. Gender-Heterogeneous Working Groups Produce Higher Quality Science. PLoS One [Internet]. 2013 Oct 30;8(10):e79147. Available from: 10.1371/journal.pone.0079147

59. Bührer S, Kalpazidou Schmidt E, Palmén R, Reidl S. Evaluating gender equality effects in research and innovation systems. Scientometrics [Internet]. 2020 Nov 1;125(2):1459–75. Available from: 10.1007/s11192-020-03596-1

60. Sebo P, Clair C. Gender Inequalities in Citations of Articles Published in High-Impact General Medical Journals: a Cross-Sectional Study. J Gen Intern Med [Internet]. 2023 Feb 1;38(3):661–6. Available from: 10.1007/s11606-022-07717-9

61. Nielsen MW, Andersen JP. Global citation inequality is on the rise. Proc Natl Acad Sci [Internet]. 2021 Feb 16 [cited 2025 Apr 24];118(7):e2012208118. Available from: 10.1073/pnas.2012208118

62. Merton RK. The Matthew Effect in Science. Science [Internet]. 1968 Jan 5 [cited 2025 Nov 28];159(3810):56–63. Available from: 10.1126/science.159.3810.56

63. Ebadi A, Schiffauerova A. How to boost scientific production? A statistical analysis of research funding and other influencing factors. Scientometrics [Internet]. 2016 Mar 1;106(3):1093–116. Available from: 10.1007/s11192-015-1825-x

64. Andersen JP, Schneider JW, Jagsi R, Nielsen MW. Gender variations in citation distributions in medicine are very small and due to self-citation and journal prestige. eLife [Internet]. 2019 July 15;8:e45374. Available from: 10.7554/eLife.45374

65. Plevris V. From Integrity to Inflation: Ethical and Unethical Citation Practices in Academic Publishing. J Acad Ethics [Internet]. 2025 Apr 21; Available from: 10.1007/s10805-025-09631-1

66. Mendoza M. Differences in Citation Patterns across Areas, Article Types and Age Groups of Researchers. Publications [Internet]. 2021;9(4):47. Available from: 10.3390/publications9040047

67. Drivas K. The evolution of order of authorship based on researchers’ age. Scientometrics [Internet]. 2024 Sept 1;129(9):5615–33. Available from: 10.1007/s11192-024-05124-x

68. Rørstad K, Aksnes DW. Publication rate expressed by age, gender and academic position – A large-scale analysis of Norwegian academic staff. J Informetr [Internet]. 2015 Apr 1;9(2):317–33. Available from: 10.1016/j.joi.2015.02.003

69. Costas R, Bordons M. Do age and professional rank influence the order of authorship in scientific publications? Some evidence from a micro-level perspective. Scientometrics [Internet]. 2011 July 1;88(1):145–61. Available from: 10.1007/s11192-011-0368-z

70. Larivière V, Pontille D, Sugimoto CR. Investigating the division of scientific labor using the Contributor Roles Taxonomy (CRediT). Quant Sci Stud [Internet]. 2021 Apr 8 [cited 2025 Nov 23];2(1):111–28. Available from: 10.1162/qss_a_00097

71. Fernandes JM, Cortez P. Alphabetic order of authors in scholarly publications: a bibliometric study for 27 scientific fields. Scientometrics [Internet]. 2020 Dec 1;125(3):2773–92. Available from: 10.1007/s11192-020-03686-0

72. Helgesson G. Authorship order and effects of changing bibliometrics practices. Res Ethics [Internet]. 2020 Jan 1 [cited 2025 Nov 27];16(1–2):1–7. Available from: 10.1177/1747016119898403

73. Győrffy B, Csuka G, Herman P, Török Á. Is there a golden age in publication activity?—an analysis of age-related scholarly performance across all scientific disciplines. Scientometrics [Internet]. 2020 Aug 1;124(2):1081–97. Available from: 10.1007/s11192-020-03501-w

74. Cammarota A, Siebenhüner AR, Olungu C, Szturz P, Güven DC, Puccini A, et al. Research training, barriers, and career development needs of early-career investigators in oncology: an EORTC survey-based study. ESMO Gastrointest Oncol [Internet]. 2025 Sept 1;9:100208. Available from: 10.1016/j.esmogo.2025.100208

75. Savage WE, Olejniczak AJ. Do senior faculty members produce fewer research publications than their younger colleagues? Evidence from Ph.D. granting institutions in the United States. Scientometrics [Internet]. 2021 June 1;126(6):4659–86. Available from: 10.1007/s11192-021-03957-4

76. Johann D, Neufeld J, Thomas K, Rathmann J, Rauhut H. The impact of researchers’ perceived pressure on their publication strategies. Res Eval [Internet]. 2024 Mar 25 [cited 2025 Nov 28];rvae011. Available from: 10.1093/reseval/rvae011

77. Gan Y, Liu J, Zhao Y, Zhu M, Wang G. Inverted U-Shaped relationship between team size and citation impact: Mediating role of responsibility diffusion. Account Res [Internet]. 2025 May 19;32(4):509–29. Available from: 10.1080/08989621.2023.2300255

78. Gonzalez-Brambila C, Veloso FM. The determinants of research output and impact: A study of Mexican researchers. Res Policy [Internet]. 2007 Sept 1;36(7):1035–51. Available from: 10.1016/j.respol.2007.03.005

79. Way SF, Morgan AC, Clauset A, Larremore DB. The misleading narrative of the canonical faculty productivity trajectory. Proc Natl Acad Sci [Internet]. 2017 Oct 31 [cited 2025 Nov 28];114(44):E9216–23. Available from: 10.1073/pnas.1702121114

80. Stevens MR, Park K, Tian G, Kim K, Ewing R. Why Do Some Articles in Planning Journals Get Cited More than Others? J Plan Educ Res [Internet]. 2022 Sept 1 [cited 2025 Nov 28];42(3):442–63. Available from: 10.1177/0739456X19827083

81. Tahamtan I, Safipour Afshar A, Ahamdzadeh K. Factors affecting number of citations: a comprehensive review of the literature. Scientometrics [Internet]. 2016 June;107(3):1195–225. Available from: 10.1007/s11192-016-1889-2

82. Sjögårde P, Didegah F. The association between topic growth and citation impact of research publications. Scientometrics [Internet]. 2022 Apr 1;127(4):1903–21. Available from: 10.1007/s11192-022-04293-x

83. Repiso R, Moreno-Delgado A, Aguaded I. Factors affecting the frequency of citation of an article. Iberoam J Sci Meas Commun [Internet]. 2021;1(1):007. Available from: https:doi.org/0.47909/ijsmc.08

84. Petersen AM, Fortunato S, Pan RK, Kaski K, Penner O, Rungi A, et al. Reputation and impact in academic careers. Proc Natl Acad Sci [Internet]. 2014 Oct 28 [cited 2025 Nov 29];111(43):15316–21. Available from: 10.1073/pnas.1323111111

85. Li W, Zhang S, Zheng Z, Cranmer SJ, Clauset A. Untangling the network effects of productivity and prominence among scientists. Nat Commun [Internet]. 2022 Aug 20;13(1):4907. Available from: 10.1038/s41467-022-32604-6

86. Vieira ES. The influence of research collaboration on citation impact: the countries in the European Innovation Scoreboard. Scientometrics [Internet]. 2023 June 1;128(6):3555–79. Available from: 10.1007/s11192-023-04715-4

87. Bunker Whittington K, King MM, Cingolani I. Structure, status, and span: gender differences in co-authorship networks across 16 region-subject pairs (2009–2013). Scientometrics [Internet]. 2024;129(1):147–79. Available from: 10.1007/s11192-023-04885-1

88. Office for National Statistics. Ethnic group, England and Wales: Census 2021 [Internet]. Newport, Wales; 2022. Available from: https://www.ons.gov.uk/peoplepopulationandcommunity/culturalidentity/ethnicity/bulletins/ethnicgroupenglandandwales/census2021

89. Higher Education Statistics Agency (HESA). Higher Education Staff Statistics: UK, 2023/24 [Internet]. Cheltenham, England; 2025 Jan. Available from: https://www.hesa.ac.uk/news/28-01-2025/sb270-higher-education-staff-statistics

90. Office for National Statistics. Annual Population Survey - Employment by occupation by sex. Survey period Jul 2024 - Jun 2025) [Internet]. Durham, England: Nomis - Official Census and Labour Market Statistics; 2025. (Employment by Occupation (SOC2020) by sex). Available from: https://www.nomisweb.co.uk/datasets/aps218/reports/employment-by-occupation?compare=E92000001

91. NHS England. NHS Workforce Statistics - June 2022 [Internet]. Leeds, England: Department of Health and Social Care; 2023 Apr. Available from: https://www.ethnicity-facts-figures.service.gov.uk/workforce-and-business/workforce-diversity/nhs-workforce/latest/

92. University of Oxford. Equality, Diversity and Inclusion Report 2023-2024 [Internet]. Oxford, England: Equality and Diversity Unit; 2025. Available from: https://edu.admin.ox.ac.uk/sites/default/files/edu/documents/media/equality_diversity_and_inclusion_report_2023_24.pdf

93. Snipe T, Jessel, K. Combined Equality Standards Report (WRES/WDES/GPG) 2024 [Internet]. Oxford: Oxford University Hospitals NHS Foundation Trust; 2024 Sept. Report No.: TB2024.77. Available from: https://www.ouh.nhs.uk/media/b2khe003/tb202477-esr-report-24.pdf

94. National Institute for Health and Care Research (NIHR). Diversity Data Report 2022. Award Funding and Selection Committees Analysis 2021-22 [Internet]. Leeds, England; 2022 Nov. Available from: https://www.nihr.ac.uk/diversity-data-report-2022

95. El Boghdady M. Equality and diversity in research: building an inclusive future. BMC Res Notes [Internet]. 2025 Jan 14;18(1):14. Available from: 10.1186/s13104-025-07096-4

96. Andersen JP, Nielsen MW, Simone NL, Lewiss RE, Jagsi R. COVID-19 medical papers have fewer women first authors than expected. eLife [Internet]. 2020 June 15;9:e58807. Available from: 10.7554/eLife.58807

97. Fulweiler RW, Davies SW, Biddle JF, Burgin AJ, Cooperdock EHG, Hanley TC, et al. Rebuild the Academy: Supporting academic mothers during COVID-19 and beyond. PLoS Biol [Internet]. 2021 Mar 9;19(3):e3001100. Available from: 10.1371/journal.pbio.3001100

98. King MM, Frederickson ME. The Pandemic Penalty: The Gendered Effects of COVID-19 on Scientific Productivity. Socius [Internet]. 2021 Jan 1 [cited 2025 Apr 24];7:23780231211006977. Available from: 10.1177/23780231211006977

99. Narayana S, Roy B, Merriam S, Yecies E, Lee RS, Mitchell JL, et al. Minding the Gap: Organizational Strategies to Promote Gender Equity in Academic Medicine During the COVID-19 Pandemic. J Gen Intern Med [Internet]. 2020 Dec 1;35(12):3681–4. Available from: 10.1007/s11606-020-06269-0

100. NIHR Oxford Biomedical Research Centre. Equality, Diversity and Inclusion Strategy 2023-2027 [Internet]. Oxford, England: Oxford University Hospitals NHS Foundation Trust; 2023 Dec. Available from: https://oxfordbrc.nihr.ac.uk/wp-content/uploads/2023/12/99657_OxBRC_Equality_Diversity_and_Inclusion_Report_2023-27_Accessible_Version.pdf

101. Oxford University Hospitals NHS Foundation Trust. Equality, Diversity, and Inclusion Objectives 2022 - 2026. Oxford, England; 2022 Sept.

102. University of Oxford. Everyone Belongs. Equality, Diversity and Inclusion Strategic Plan 2024-2027 [Internet]. Oxford, England; 2024 Sept. Available from: https://edu.admin.ox.ac.uk/sites/default/files/edu/documents/media/oxford_university_edi_strategic_plan_2024_27.pdf

103. Boulware LE, Giselle C, Aguilar-Gaxiola S, Wilkins CH, Ruiz, Vitale A, et al. Combating Structural Inequities — Diversity, Equity, and Inclusion in Clinical and Translational Research. N Engl J Med [Internet]. 2022 Jan 19 [cited 2025 Apr 24];386(3):201–3. Available from: 10.1056/NEJMp2112233

104. Hinton A, Lambert WM. Commentary: Moving diversity, equity, and inclusion from opinion to evidence. Cell Rep Med [Internet]. 2022 Apr 19;3(4):100619. Available from: https://www.sciencedirect.com/science/article/pii/S2666379122001367

105. Shah SS. Rapid Response: Need for addressing structural and cultural barriers to equality, diversity and inclusion in the NIHR BRCs [Re: Effect of Athena SWAN funding incentives on women’s research leadership]. BMJ [Internet]. 2021 Jan 24 [cited 2022 Oct 3];371:3975/rr. Available from: https://www.bmj.com/content/371/bmj.m3975/rr

